# Estimating the Treatment Effects of Multiple Drug Combinations on Multiple Outcomes in Hypertension

**DOI:** 10.1101/2024.11.10.24317054

**Authors:** Ruoqi Liu, Lang Li, Ping Zhang

**Author notes:** Corresponding Author & Lead Contact.

## Abstract

Hypertension poses a significant global health challenge, and its management is often complicated by the complexity of treatment strategies involving multiple drug combinations and the need to consider multiple outcomes. Traditional treatment effect estimation (TEE) methods struggle to address this complexity, as they typically focus on binary treatments and binary outcomes. To overcome these limitations, we introduce **METO**, a novel framework designed for TEE in the context of multiple drug combinations and multiple outcomes. **METO** employs a multi-treatment encoding mechanism to handle multiple drug combinations and their sequences effectively, and differentiates between effectiveness and safety outcomes by explicitly learning the outcome type when predicting the treatment outcomes. Furthermore, to address confounding bias in outcome prediction, we employ an inverse probability weighting method tailored for multiple treatments, assigning each patient a balance weight derived from their propensity score against different drug combinations. Our comprehensive evaluation using a real-world patient dataset demonstrates that **METO** outperforms existing TEE methods, with an average improvement of 5.0% in area under the precision-recall curve and 6.4% in influence function-based precision of estimating heterogeneous effects. A case study demonstrates that our method successfully identifies personalized optimal antihypertensive dual regimens, achieving maximal efficacy and minimal drug-related safety risks. This showcases its potential for improving treatment strategies and outcomes in hypertension management.

## Introduction

Hypertension is a major global health issue, responsible for around 7 million deaths and 57 million disabilities annually ^1^. However, optimal blood pressure control remains elusive for roughly 70% of patients, primarily due to inadequate implementation of combination therapies ^2^. The challenge lies in selecting effective antihypertensive treatment strategies ^3^, including the choice between starting with monotherapy and gradually adding another drug (stepped-care) or beginning with a drug combination ^4,5^.

This introduces a pivotal medical question: “*How can we determine the most effective antihypertensive drug combinations (treatments) to improve hypertension-related conditions (outcomes)?”* Tackling this question requires an exploration into the complex landscape of treatment options, encompassing a variety of drug combinations and sequences. Moreover, the imperative for a thorough evaluation of treatments underscores the inherent tension between effectiveness and safety outcomes in hypertension ^6^, illuminating the dual, often opposing, dimensions of outcome assessment ^7^.

Treatment effect estimation (TEE), which identifies the causal effects of *treatments* on the patient *outcomes*, can be leveraged to address the above complex question of optimizing antihypertensive drug combinations. However, existing TEE approaches, designed predominantly for binary treatments and binary outcomes ^8–10^, struggle with multiple treatments and multiple outcomes. Some approaches ^11–14^ extend existing models for multiple treatments by directly increasing treatment arms, thus facing efficiency and generalizability challenges ^15^. Others improve adaptability using unified treatment embeddings for multiple treatments ^15,16^. Nevertheless, significant challenges remain in applying these methods to TEE with multiple treatments and multiple outcomes, particularly for antihypertensive drug combinations.

First, existing methods ^13,16^ typically treat all treatments uniformly, without utilizing the nuanced details of each. This lack of granular differentiation limits their practicality, especially in the studied scenarios where the combination and sequential administration of drugs are crucial. Second, while some studies ^17–19^ have explored TEE in the context of multiple outcomes, they are mainly designed for randomized controlled trials without adjustment for confounding bias, and more importantly, fail to differentiate between outcome types (i.e., therapeutic effectiveness versus adverse effects). In addition, different outcomes can have different relationships with the covariates and treatments. Failing to consider such various relationships may lead to confounding bias and inaccurate treatment effects estimation. Finally, there is a noticeable gap in the literature regarding the comprehensive treatment effect assessment tool that facilitates clinical decision-making.

To address these challenges, we propose a novel framework, called **METO**, to estimate the treatment effects of **M**ultipl**E** drug combinations on mul**T**iple **O**utcomes for identifying optimal antihypertensive drug combinations (see Fig. 1). First, we address the complexities of multiple drug combinations through the proposed multi-treatment modeling. This mechanism processes the detailed information of drug combinations and administration sequences independently, before synthesizing these elements via a deep fusion layer. Second, we leverage the specific outcome type information as additional guidance to differentiate between effectiveness and safety outcomes, enhancing the accuracy of outcome prediction. In addition, to address confounding bias in outcome prediction, we employ an inverse probability weighting method tailored for multiple treatments, assigning each patient a balance weight derived from their propensity score against different drug combinations. Our comprehensive evaluation shows that METO outperforms existing treatment effect estimation methods and successfully identifies personalized optimal drug combinations with beneficial effects and reduced safety risks.

**Figure 1:**
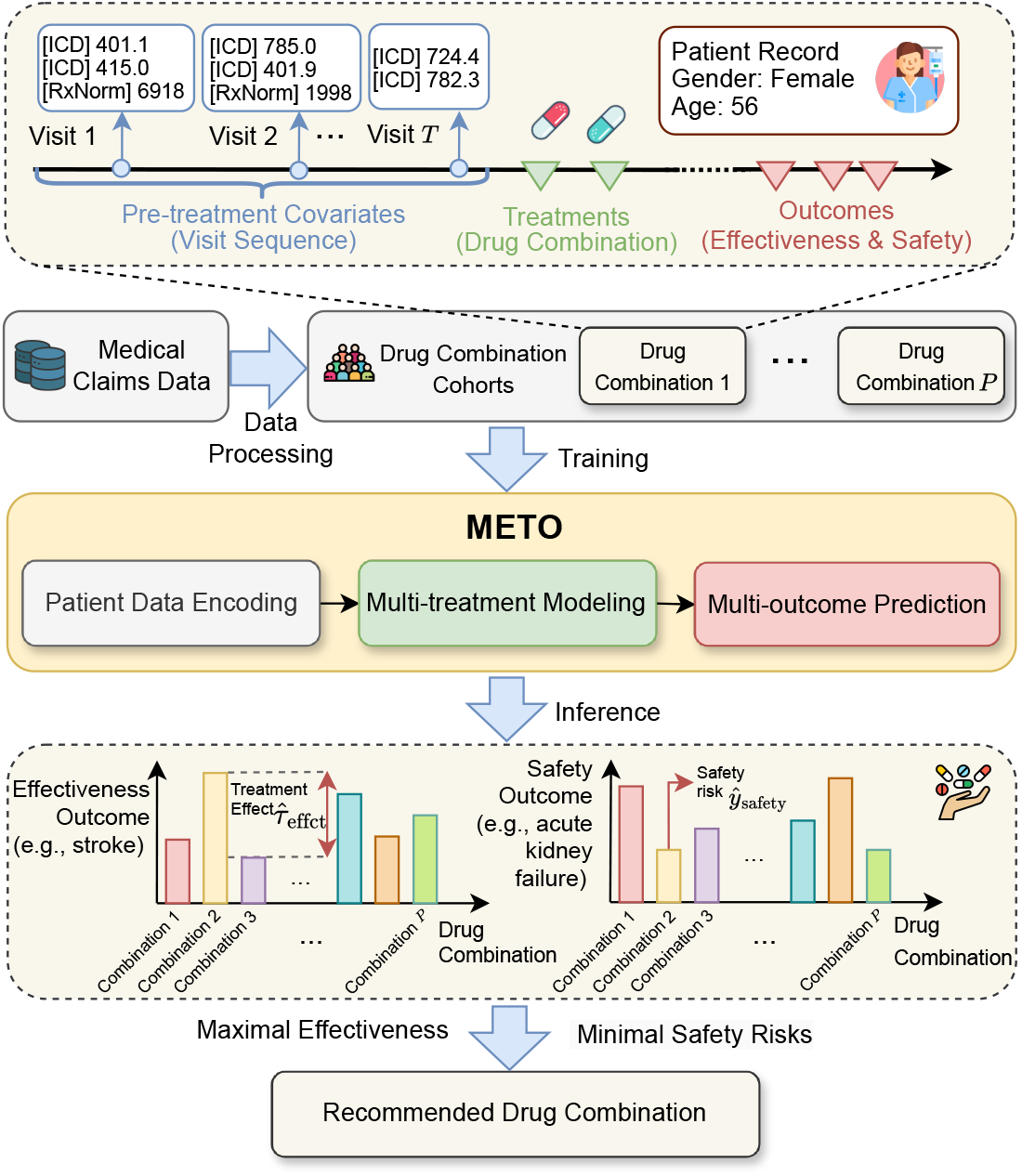
Workflow overview of the proposed **METO**. Patient records are extracted from medical claims data and processed to train the model, which learns to estimate the treatment effects of multiple drug combinations. The lower panel shows the model’s output, where the estimated effects on both effectiveness and safety outcomes assist clinicians in making informed treatment decisions for hypertension.

Our contributions are summarized as follows:

- **Problem**. We address the challenge of TEE in hypertension management, focusing on multiple drug combinations and their impact on both effectiveness and safety outcomes.
- **Method**. We propose **METO**, a novel method that incorporates multi-treatment encoding and explicit outcome-type learning to handle complex drug combinations and differentiate between effectiveness and safety outcomes.
- **Experiments**. We validate **METO**’s superior performance against existing TEE methods using a largescale real-world dataset and demonstrate its practical efficacy through a case study on personalized drug combination recommendation.

## Results

### Overall Framework

We develop an end-to-end treatment effect estimation framework that can be utilized for recommending optimal drug combinations for patients with hypertension. As shown in Fig. 1, the patient medical records are extracted from an observational database and then processed into drug combination cohorts for comparing treatment effectiveness. The treatment effects are estimated with the proposed **METO**, which is designed specifically for the scenario of multiple treatments and multiple outcomes. Finally, optimal drug combinations are suggested by assessing the treatment effects across both effectiveness and safety outcomes.

### Dataset

We collect data on approximately 130 million patients from the MarketScan Commercial Claims and Encounters (CCAE)^1^ database, covering the period from 2012 to 2021. This dataset contains individual-level, de-identified healthcare claims information from employers, health plans, and hospitals. We extract more than 19 million patients who are diagnosed with hypertension (Appendix Table A1). Within this patient cohort, we identify more than 11 million patients who have at least one risk factor (i.e., subclinical organ damage, diabetes, renal, or associated cardiovascular disease) ^20^ and initiated first-line antihypertensive drugs. Each patient record consists of demographics (e.g., age and sex), co-morbidities (via ICD-9/10 diagnosis codes) as well as co-prescribed medications (using national drug codes [NDC]). For uniformity, ICD-9/10 codes were consolidated into a standardized coding schema using the clinical classifications software (CCS)^2^. Figure 2 displays the dataset’s statistics including the distribution of overall population, age, gender, and outcome across different drug combinations, respectively. Figure 3 illustrates the detailed study design for each treatment cohort, including treatment definitions, computation of confounding variables, and outcomes. The flowchart of the user cohort selection is provided in Appendix Fig. A1.

**Figure 2:**
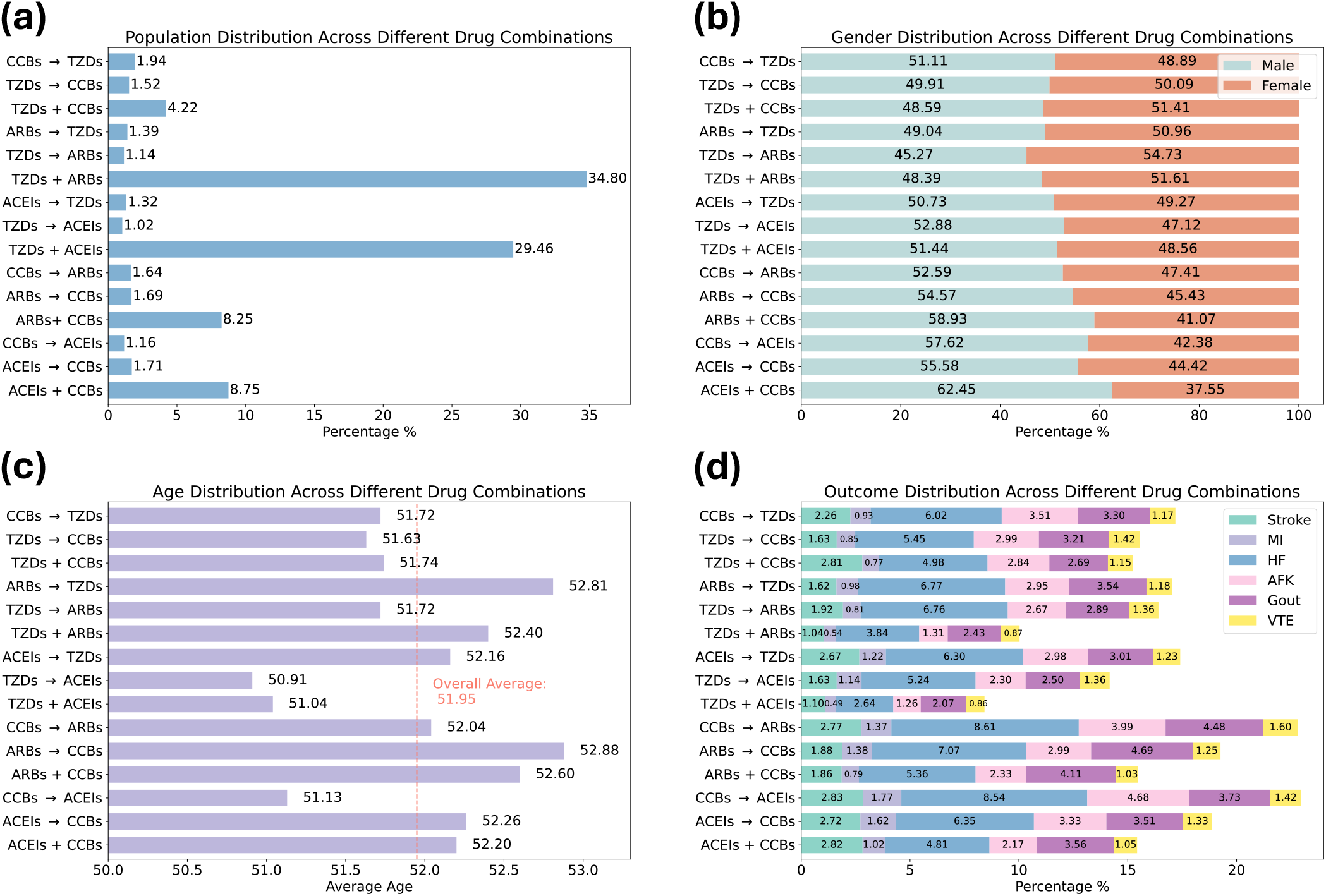
Statistics of hypertension patient cohorts across 15 unique drug combinations. (a) Patient population distribution across different drug combinations. (b) Gender distribution across different drug combinations. (c) Age distribution across different drug combinations. (d) Outcome distribution across different drug combinations. TZDs: thiazide diuretics. ACEIs: ACE inhibitors. ARBs: angiotensin receptor blockers. CCBs: calcium channel blockers. + and → denote the initial combination and stepped-care, respectively (e.g., “ACEIs + CCBs” represents the initial combination of ACEIs and CCBs, while “ACEIs → CCBs” represents first ACEIs, then CCBs sequentially). MI: acute myocardial infarction. HF: heart failure. AFK: acute kidney failure. VTE: venous thromboembolism

**Figure 3:**
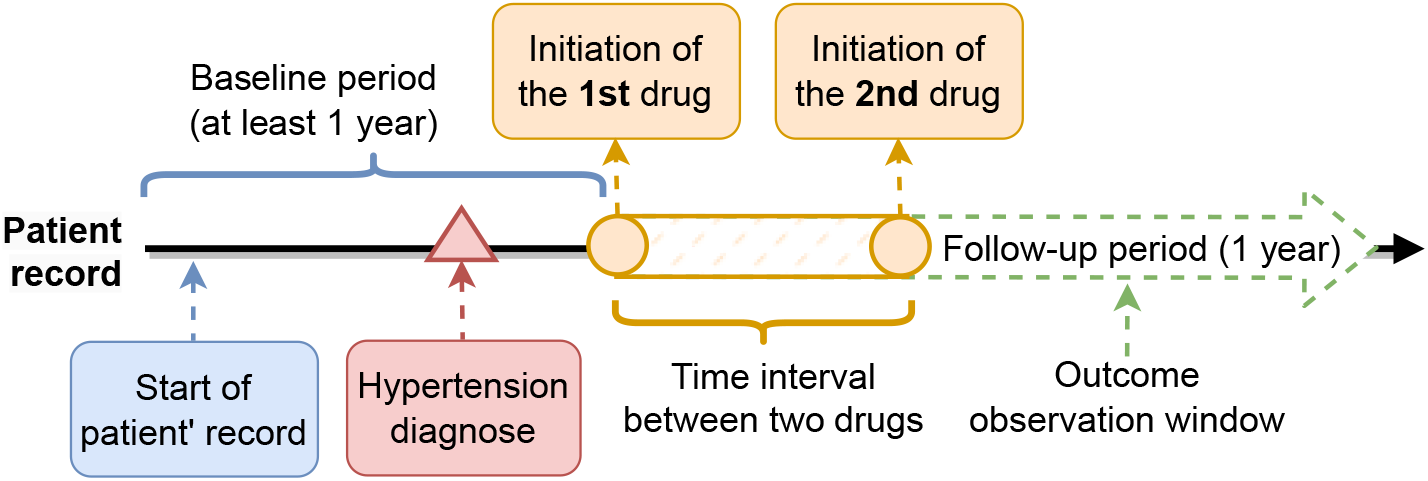
Illustratioin of the treatment cohort definitions. The treatment consists of two drugs as a combination. The outcomes are computed in the follow-up period. The pre-treatment covariates obtained from the baseline period are regarded as confounders for adjustment.

### Treatments

Following hypertension management guidelines ^3^, we categorized first-line antihypertensive agents into thiazide diuretics (TZDs), ACE inhibitors (ACEIs), angiotensin receptor blockers (ARBs), and calcium channel blockers (CCBs). We identified five distinct first-line drug combination regimens, each comprising two different classes: (1) TZDs and ACEIs; (2) TZDs and ARBs; (3) TZDs and CCBs; (4) ACEIs and CCBs; (5) ARBs and CCBs. We note that ACEIs and ARBs are not combined in clinical practice. These combinations also varied by assignment order: initial combination (less than 30 days between first and second drug initiations) and stepped-care (between 30 and 180 days). We identified a total of 15 unique drug combinations, considering both the specific combinations and their order of assignment. Detailed definitions of these drugs are provided in Appendix Table A2.

### Outcomes

Six important outcomes are computed during the follow-up period after the treatment initialization, including three primary effectiveness outcomes (stroke, acute myocardial infarction [MI], heart failure [HF]) and three safety outcomes (acute kidney failure [AKF], gout, venous thromboembolism [VTE]). These outcomes are chosen based on their significance in hypertension management guidelines ^3^ and insights from recent large-scale studies in hypertension ^6^. Outcome occurrences are identified using diagnosis codes, with detailed definitions available in Appendix Table A3.

### Confounders

A comprehensive list of potential confounders is compiled, including demographics (age and gender), 282 unique co-morbidities (based on diagnosis codes), and 1,378 unique co-prescribed medications. These confounders, relevant to both treatment assignment and outcomes, are assessed during the baseline period before treatment initiation.

### Baselines and Setup

In the experiments, we compare our method with state-of-the-art baselines, which can be classified into three main categories:

- *Basic meta-learners*: (1) **S-learner** is a meta-learner ^21^ that builds a binary outcome prediction model for all treatment groups; (2) **T-learner** is also a meta-learner ^21^ that builds multiple outcome prediction models for each treatment group separately.
- *TEE methods for binary treatments and binary outcomes*: (1) **TARNet** ^8^ predicts the potential outcomes based on balanced representations between treated and controlled groups; (2) **DragonNet** ^9^ jointly predicts treatment and outcomes based on the shared representations via a three-head neural network.
- *TEE methods for multiple treatments and binary (multiple) outcomes*: (1) **PerfectMatch** ^11^ augments samples within a batch with their propensity-matched nearest neighbors. The framework can be naturally extended to multiple treatments by matching with the nearest neighbors from each treatment group; (2) **TECE-VAE** ^16^ incorporates latent variables and causal structure through a variational autoencoder, and models multiple treatments with a task embedding; (3) **MEMENTO** ^13^ is a direct extension of TARNet ^8^ to multiple treatments by increasing the number of model branches to the number of treatments; (4) **LR-learner** ^22^ adopts a neural network to learn embeddings for individuals and treatments, which are then transformed by a linear operator to predict outcomes based on the dot product of these embeddings; (5) **TransTEE** ^15^ embeds multiple treatments and covariates into a shared hidden space via a Transformer to improve the model’s flexibility and robustness; (6) **NCoRE** ^12^ models the cross-treatment interactions by encoding each treatment arm separately and then applying a merge layer to connect all the treatment arms.

Note that we implement the S-learner and T-learner with a logistic regression (LR)-based estimator. TARNet ^8^ and DragonNet ^9^ are representative works in TEE but initially designed for binary treatment scenarios. We extend these two methods to multiple treatment scenarios as done in ^11^. For baselines originally developed for binary outcomes, we have facilitated their application to multiple outcome contexts by transitioning from a binary classification head to one capable of multi-label classification.

### Evaluation Metrics

We evaluate the performance of factual outcome prediction by measuring the area under the Precision-Recall curve (AUPR), focusing on the precision-recall trade-off due to the potential imbalance between positive and negative outcome labels. As the ground truth treatment effects are not observed (i.e., any patient is ever only assigned to one of the treatments in practice), we can not directly compute the traditional metric precision in estimating heterogeneous effects (PEHE) ^23^. Instead, we adopt a recent proxy metric for counterfactual evaluation, called influence function-based precision of estimating heterogeneous effects (IF-PEHE) ^24^, which measures the mean squared error between estimated treatment effects and approximated true treatment effects (see detailed computation of IF-PEHE [IP] in Appendix C.2). The original metric is designed for binary treatments and binary outcomes, we extend it to multiple treatments and multiple outcomes (MM-IP) as below:

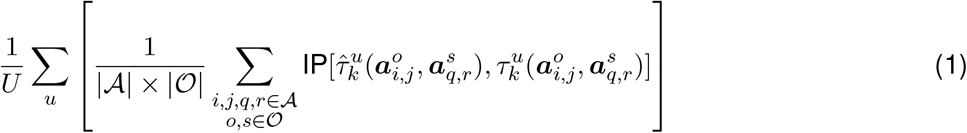

where U is the total number of patients, 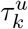 denotes the treatment effect for patient u on outcome k.

### Implementation Details

The dataset is randomly split into training, validation, and test sets with percentages of 80%, 10%, and 10% respectively. The number of training epochs is 10 and the learning rate is 5e-5. The backbone model architecture is a 12-layer Transformer, with 768 hidden units, 12 attention heads, and 3072 for the intermediate size. The parameter *β* in multi-treatment modeling is set to 0.6. The parameter *η* in outcome type-informed prediction is set to 1 for the training and 0 for the inference. The parameter *λ*, which adjusts the influence of treatment prediction, is set to 1. All results are reported on the test sets over 20 initializations of the model. More implementation details including the parameter tuning, training setup, and additional configurations are mentioned in Appendix C.3.

### Overall Performance Comparison

Results in Table 1 present a comparative evaluation of our proposed **METO**’s performance against baselines in factual outcome prediction and TEE on the real-world hypertension dataset. Notably, **METO** achieves superior performance over the best baseline, registering an average improvement of 5.0% in AUPR and 6.4% in MM-IP.

**Table 1:** Performance comparison for factual outcome prediction (AUPR) and treatment effect estimation (MM-IP: IF-PEHE for multiple treatments and multiple outcomes). The results are averaged over 20 random runs. Full results with standard deviations for all methods are provided in Appendix Table A6

| Method | Stroke |  | MI |  | HF |  | AKF |  | Gout |  | VTE |  |
| --- | --- | --- | --- | --- | --- | --- | --- | --- | --- | --- | --- | --- |
|  | AUPR ↑ | MM-IP ↓ | AUPR ↑ | MM-IP ↓ | AUPR ↑ | MM-IP ↓ | AUPR ↑ | MM-IP ↓ | AUPR ↑ | MM-IP ↓ | AUPR ↑ | MM-IP ↓ |
| S-learner | 0.551 | 0.284 | 0.188 | 0.355 | 0.329 | 0.378 | 0.218 | 0.370 | 0.339 | 0.325 | 0.274 | 0.356 |
| T-learner | 0.544 | 0.297 | 0.181 | 0.360 | 0.325 | 0.386 | 0.212 | 0.377 | 0.331 | 0.332 | 0.267 | 0.363 |
| TARNet | 0.569 | 0.271 | 0.196 | 0.351 | 0.342 | 0.357 | 0.233 | 0.359 | 0.349 | 0.318 | 0.288 | 0.355 |
| DragonNet | 0.574 | 0.262 | 0.198 | 0.344 | 0.351 | 0.338 | 0.251 | 0.342 | 0.356 | 0.303 | 0.294 | 0.332 |
| PerfectMatch | 0.591 | 0.239 | 0.202 | 0.361 | 0.388 | 0.312 | 0.274 | 0.331 | 0.378 | 0.289 | 0.312 | 0.317 |
| MEMENTO | 0.584 | 0.244 | 0.208 | 0.354 | 0.382 | 0.317 | 0.270 | 0.338 | 0.372 | 0.294 | 0.310 | 0.322 |
| TECE-VAE | 0.605 | 0.230 | 0.211 | 0.355 | 0.397 | 0.302 | 0.290 | 0.325 | 0.381 | 0.283 | 0.319 | 0.309 |
| LR-learner | 0.609 | 0.225 | 0.206 | 0.364 | 0.392 | 0.308 | 0.296 | 0.321 | 0.385 | 0.280 | 0.326 | 0.315 |
| TransTEE | 0.617 | 0.221 | 0.235 | 0.321 | 0.409 | 0.284 | 0.303 | 0.311 | 0.403 | 0.261 | 0.338 | 0.276 |
| NCoRE | 0.577 | 0.249 | 0.207 | 0.361 | 0.388 | 0.313 | 0.275 | 0.334 | 0.370 | 0.297 | 0.311 | 0.326 |
| **METO** | **0.656** | **0.185** | **0.329** | **0.225** | **0.442** | **0.236** | **0.364** | **0.238** | **0.453** | **0.204** | **0.363** | **0.201** |
| (std) | 0.011 | 0.003 | 0.009 | 0.005 | 0.013 | 0.004 | 0.014 | 0.007 | 0.007 | 0.006 | 0.008 | 0.003 |

The basic meta-learners, namely the S-learner and T-learner, exhibit diminished performance in comparison to advanced deep learning-based TEE approaches. This discrepancy is attributed to their inability to adequately process high-dimensional, heterogeneous patient data and to forge precise patient representations for accurate effect estimation. While deep learning-based methods for binary treatment and outcome scenarios, such as TARNet, show marginal enhancements over meta-learners, they fall short in addressing the intricacies of multiple treatments and outcomes.

Methods designed for multiple treatments, like PerfectMatch and MEMENTO, offer advancements over binary treatment and outcome frameworks. However, their rigid model architectures limit their adaptability and generalizability across diverse treatment interactions, rendering their performance inferior to more flexible approaches like TECE-VAE, LR-learner, and TransTEE, which are tailored for complex treatment and outcome mappings.

The exceptional efficacy of **METO** is attributed to two main strategies: Firstly, the incorporation of a multi-treatment encoding module for the explicit encoding of multiple treatments, including the specifics of drug combinations and their administration sequencing. This approach is finely tuned to address the real-world complexities encountered with antihypertensive drugs and their combinatory uses. Secondly, **METO** distinctively models varying types of outcomes—distinguishing therapeutic effectiveness from safety outcomes through the proposed outcome type-informed prediction. By acknowledging the inherent differences in their practical implications and the variability in their distribution across the population, our method adeptly navigates the dual objectives of optimizing disease progression while mitigating safety risks.

### Population Outcome Comparison

We hypothesize that the drug combination recommended by our model based on the estimated treatment effects can effectively prevent the patients from developing severe disease outcomes. To demonstrate this, we compare the prevalence of different disease outcomes against two patient treatment groups: 1) model-recommended treatment and 2) actual treatment (different from the model’s recommendation). Specifically, we first obtain a group of patients whose actual treatment is different from the model’s recommendation. Then we derive a comparison group by involving the most similar patients whose actual treatment matches the model recommendation. We use the baseline patient representations to calculate the similarity via Euclidean distance. Finally, we compute and compare the prevalence of each disease outcome in Fig. 4. We observe that, for a given outcome, the prevalence rate in patients who receive treatments that are different from our recommendation is higher than the average rate baseline, while the prevalence rate of patients who receive the same treatments as our recommendation is lower than the baseline. This illustrates that our model recommends effective treatment strategies (reflecting on lower prevalence rate on all outcomes), and provides potential clinical insights for doctors to decide the optimal drug combinations for patients with hypertension.

**Figure 4:**
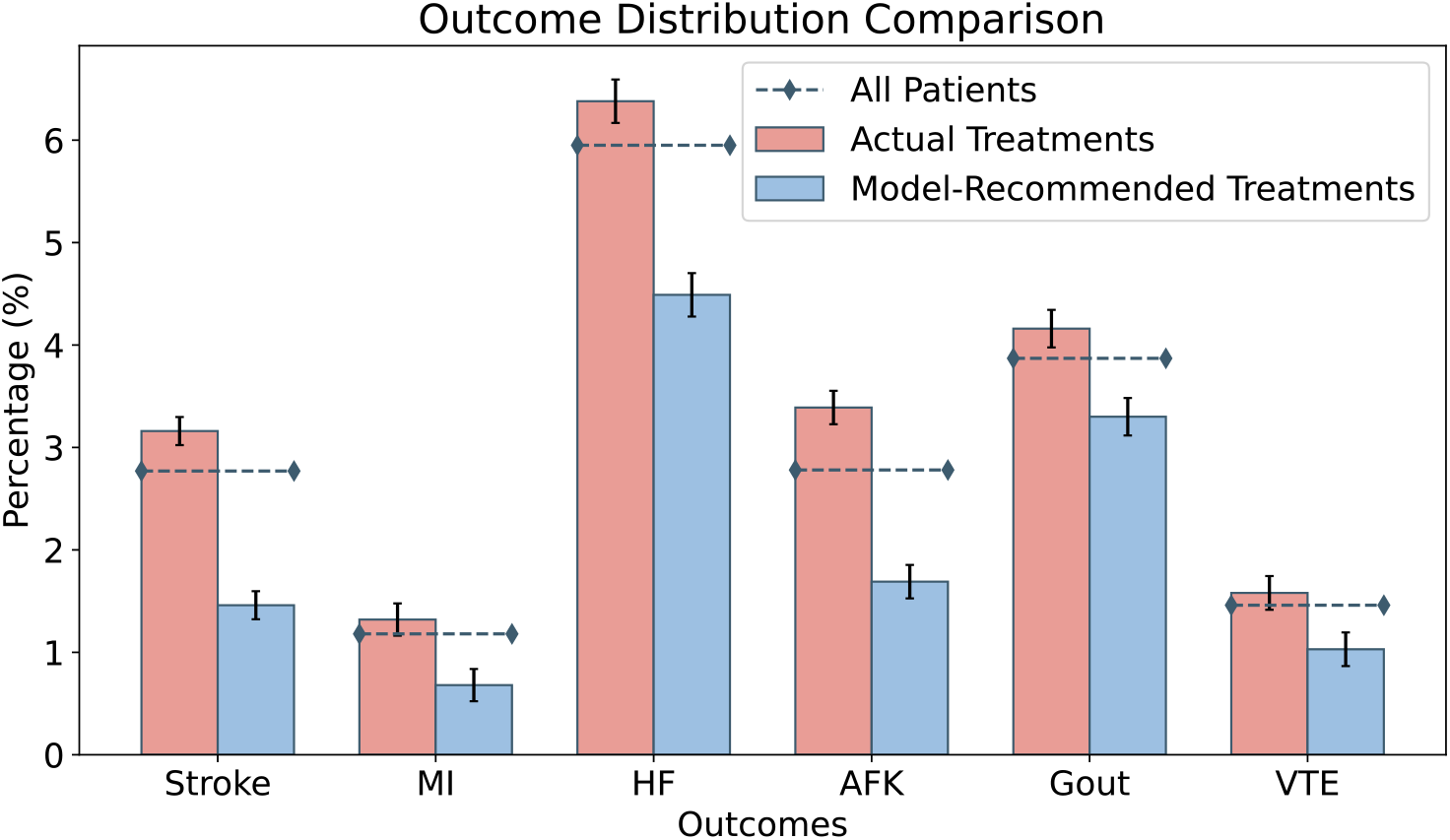
Comparison of disease outcomes for different treatment approaches among the patient population. The percentage of patients experiencing specific outcomes across three categories: all patients (dashed line with diamond markers), actual treatments administered (red bars), and model-recommended treatments (blue bars). The error bars on each column indicate the standard errors of the data points.

### Ablation Study

To demonstrate the significance of individual components within our model, we conducted a comprehensive ablation study. We explore the performance impact of various model configurations, including (1) **w/o MT**, where the multi-treatment (MT) encoding module is substituted with a generic embedding layer; (2) **w/o TC-A**, which replaces the treatment-covariate co-attention (TC-CoA) with an average pooling approach; (3) **w/o MT and TC-A**, which removes both the MT encoding and TC-CoA to understand the combined effect of these two components; (4) **w/o OT**, omitting the informative outcome types (OT) in multi-outcome prediction; (5) **w/o OC-A**, while replaces the outcome-distinctive covariate attention (OC-A) with an average pooling layer. (6) **w/o OT and OC-A** removes both the OT information and TC-CoA.

The results in Fig. 5 present model superiority across various configurations. Notably, the **w/o MT and TC-A** variant exhibits the most significant performance drop, underscoring the critical role of multitreatment encoding and treatment-covariate co-attention in effectively capturing the nuanced relationships between treatments and covariates. Similarly, the **w/o OT and OC-A** configuration shows a marked decrease in performance, highlighting the value of incorporating outcome types and the outcome-distinctive covariate attention mechanism for enhanced prediction accuracy.

**Figure 5:**
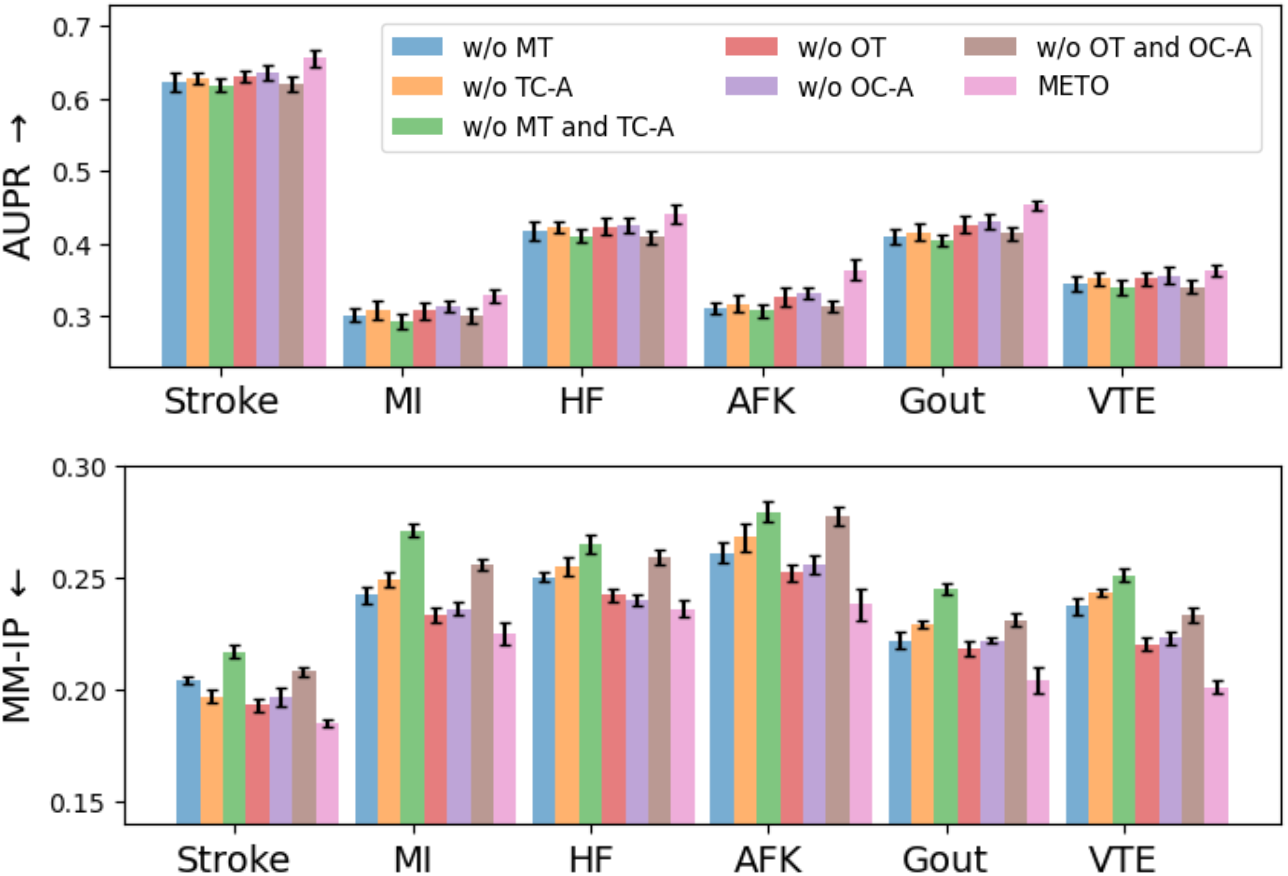
Ablation study for different variants of **METO**. The error bars on each column indicate the standard errors of the data points.

Further comparisons with nuanced model variants reveal the significance of each individual component to model performance. For example, **w/o TC-A, w/o OC-A**, which employ average pooling instead of the specialized attention mechanisms, reveal that our proposed attention-based approach is more adept at modeling the intricate interplay among covariates, treatments, and outcomes. This finding affirms the attention mechanism’s capability to refine TEE by accurately encoding complex relationships within the data. This ablation study not only confirms the integral role of each proposed component in bolstering model performance but also emphasizes the architecture’s holistic design in addressing the challenges of TEE with multiple treatments and multiple outcomes. Additional ablation study on propensity score weighting is provided in Appendix Table A7.

### Case Study

#### Treatment Recommendation

Identifying optimal antihypertension drug combinations is inherently complex due to the multiple available treatments, various drug combinations, multiple patient outcomes, and patient heterogeneity. Our model addresses this complexity by providing a comprehensive clinical outcome assessment tool that evaluates the effects of various drug combinations on both therapeutic effectiveness and safety outcomes. Through a detailed case study depicted in Fig. 6, we demonstrate the practical application of our model in guiding antihypertensive treatment decisions. Within this framework, a patient’s medical record is analyzed to estimate the effects of all potential drug combinations. These estimates are then prioritized according to their effectiveness, with a simultaneous assessment of associated safety risks, providing a comprehensive overview for clinical review. In the illustrated case, the model identified the combination of TZDs and ACEIs initiated simultaneously as the strategy offering the most significant therapeutic benefit and minimum safety concerns. This finding exemplifies the model’s ability to recommend treatment plans that optimize for both efficacy and safety, presenting a valuable assessment and decision-support tool for clinicians.

**Figure 6:**
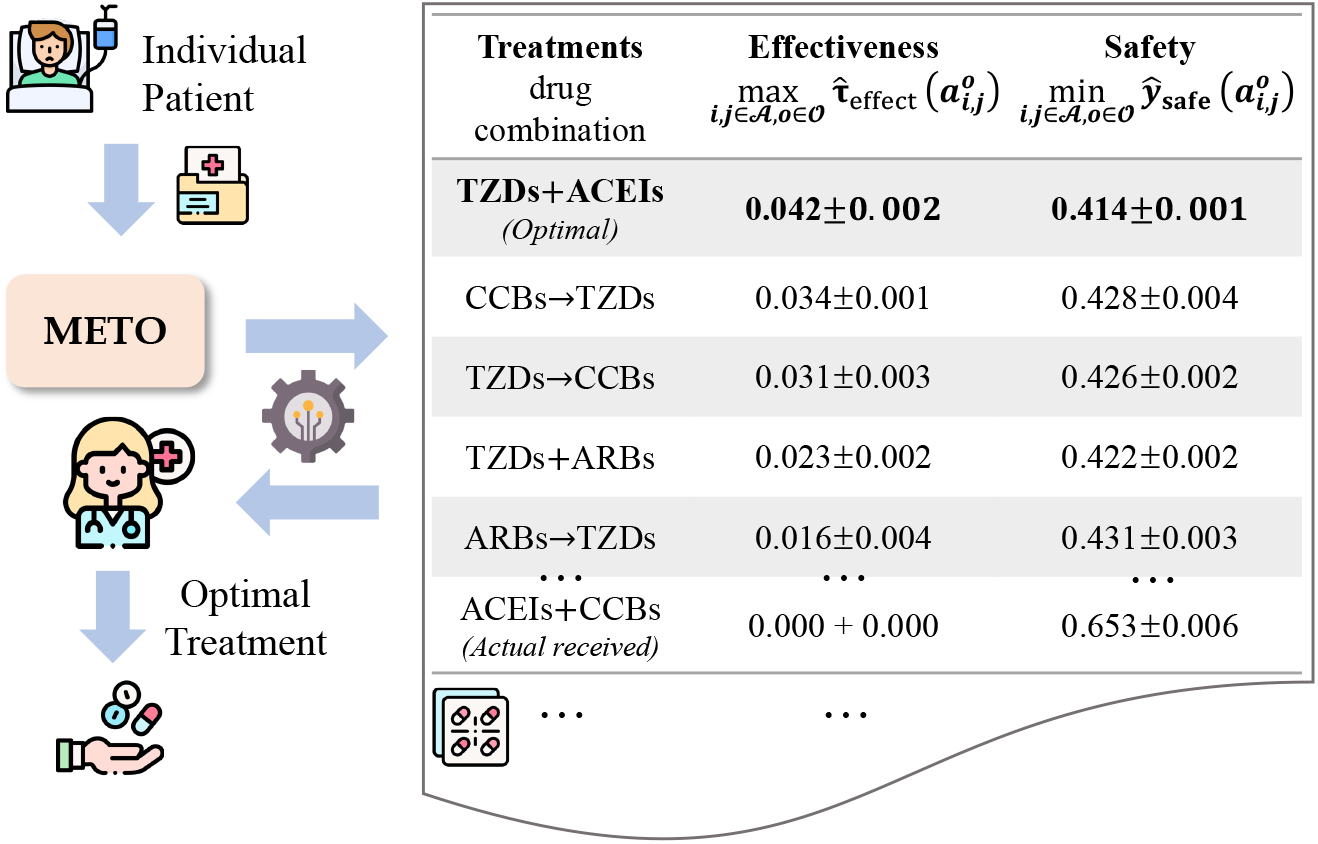
Illustration of how the proposed **METO** can be used to assist clinicians decide the personalized optimal drug combination with beneficial effects and reduced safety risks.

#### Attention Visualization

For investigating the effect of outcome-distinctive covariate attention, we visualize the attention weights on the patient pre-treatment covariates using a heatmap in Fig. 7. It presents the top 20 covariates ranked by the learned attention weights of a patient who was prescribed ACEIs and CCBs as initial combination therapy and subsequently developed HF and VTE during their disease progression. This heatmap visualization elucidates the distinct and shared covariate relevancies across the two outcome categories, highlighting how certain conditions such as “respiratory failure; insufficiency; arrest” and “pulmonary heart disease” have high relevance for both HF and VTE. Conversely, specific conditions like “coagulation and hemorrhagic disorders” show a pronounced association primarily with VTE, reflecting the model’s nuanced understanding of different mechanisms of effectiveness and safety outcomes. This analysis confirms the capability of the proposed attention networks to dynamically concentrate on both unique and shared information pertinent to patient outcomes. By effectively capturing and modeling the intricate relationships between various covariates and outcomes, attention mechanisms play a crucial role in enhancing the model’s predictive accuracy in TEE, thereby supporting personalized treatment strategies.

**Figure 7:**
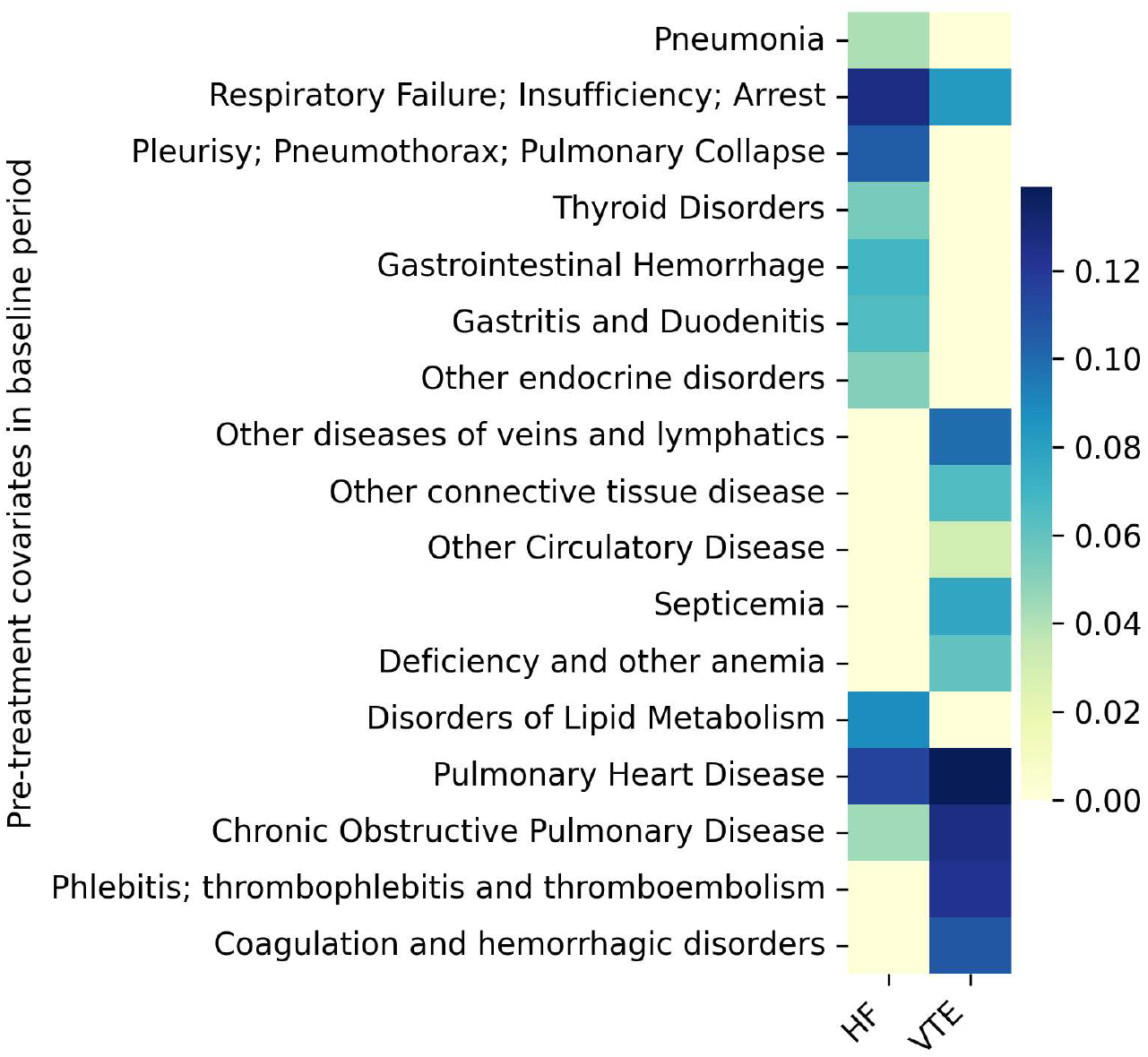
Visualization of covariates with largest outcome-distinctive covariate attention (*α*_*k*_) of one patient with two outcomes: HF and VTE.

## Discussion

In this paper, we studied the problem of treatment effect estimation in the complex context of hypertension management, characterized by multiple drug combinations and multiple outcomes. We proposed **METO**, an innovative methodology designed to address these complexities. **METO** employs multi-treatment encoding to unravel the intricacies of various drug combinations and leverages outcome type information to better differentiate between effectiveness and safety outcomes. Validated by extensive experiments on a real-world dataset, **METO** demonstrates superior performance compared to traditional TEE methods. By offering a more comprehensive treatment effect assessment for antihypertensive drugs, **METO** makes a substantial contribution to improving patient care in the field of hypertension management.

### TEE under Multiple Treatments

Traditional TEE research is largely anchored in binary treatment and binary outcome scenarios^8–10,25–28^, posing challenges when extending to accommodate the complexity of real-world healthcare scenarios involving multiple treatments and multiple outcomes. Though some studies ^11–13^ have been proposed to specifically address the challenges of multiple treatments, they often face computational inefficiencies and less flexibility. For instance, MEMENTO ^13^, which is a direct extension of a traditional TEE method (TARNet ^8^), assigns covariates from different treatment groups to different branches in their model. This approach can be less flexible with varying numbers of treatments. Methods^11–13^ that encode multiple treatments into a hidden embedding space offer some improvements in modeling flexibility. TECE-VAE ^16^, for instance, incorporates latent variables and causal structure through a variational autoencoder (VAE), and models multiple treatments with a task embedding. However, all these methods still fall short of fully capturing the complexities of multiple treatments in real applications, where the treatment can be a drug combination with different administration sequences, thus leading to an insufficient understanding of treatments and suboptimal model performance.

### TEE under Multiple Outcomes

Recent approaches^17–19,22,29^ have attempted to address TEE under multiple outcome scenarios. For example, Kennedy et al. ^17^ propose to translate the treatment effects of multiple outcomes to a common scale and then estimate these scaled effects with non-parametric statistical methods. Argaw et al. ^19^ examine the heterogeneous treatment effects and identify patient subpopulations under multiple outcomes. However, most methods are designed and evaluated on randomized controlled trial data (e.g., A/B testing) without the consideration of confounding bias (i.e., non-randomized treatment assignment) that widely exists in observational patient data. More importantly, a crucial aspect in our context is the differentiation between types of outcomes: therapeutic effectiveness versus safety endpoints. The omission of this distinction in current methodologies can lead to inaccuracies in outcome prediction and failure to identify optimal treatment strategies that consider both aspects.

### Covariate Balancing Assessment

In observational studies, it is crucial to ensure that the treatment groups are comparable with respect to baseline characteristics. We achieve this through propensity weighting, which reduces potential confounding by re-weighting each individual based on the propensity score. We compare the covariate balancing of the unweighted (original) and weighted data with absolute standard mean difference (ASMD). The ASMD is a widely used metric in propensity score analysis, quantifying the difference in means (or proportions) of a covariate between treatment groups, standardized by the pooled standard deviation. An ASMD of 0.1 or less is generally considered to indicate adequate balance. We follow existing work ^30^ to calculate the ASMD of a target treatment against the remaining treatments. Figure 8 presents the ASMD of drug combination cohort “TZDs + ACEIs” for a variety of covariates, including demographic factors (e.g., age), clinical diagnoses (e.g., thyroid disorders, diabetes mellitus with complications), and medications (e.g., furosemide, metformin). In the unweighted original data, several covariates exhibit ASMDs greater than the 0.1 threshold, indicating a significant imbalance. After re-weighting the population, the ASMDs for most covariates are reduced and fall below the 0.1 threshold, demonstrating improved balance. This enhanced balance demonstrates that the confounding bias is mitigated, ensuring the accuracy of estimated treatment effects. A comprehensive covariate balancing plot for all covariates is provided in Appendix Fig. A2.

**Figure 8:**
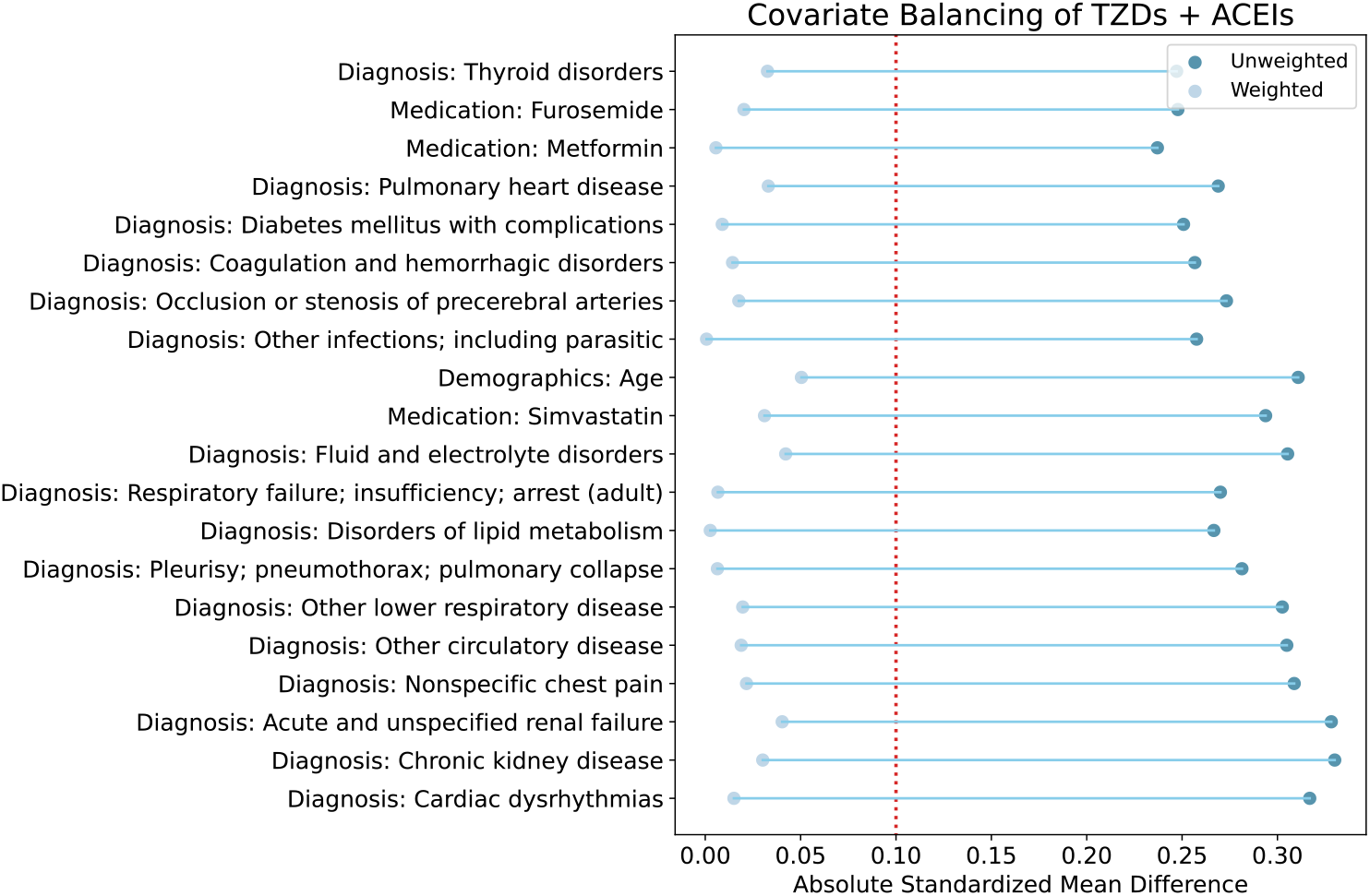
Performance of covariate balancing of TZDs and ACEIs. The absolute standard mean difference (ASMD) values of the top 20 well-balanced covariates before and after weighting are presented. The vertical dotted red line at 0.10 represents the threshold for balance, with points closer to zero indicating better balance.

### Results on Semi-Synthetic Data

Since ground truth treatment effects are unavailable in observational data (i.e., only one of the potential outcomes can be observed), direct evaluation of model performance for estimation accuracy is not possible. To address this limitation, we conducted experiments on semisynthetic data with simulated true treatment effects. Specifically, treatment assignments and outcomes were simulated based on real patient covariates (details provided in Appendix D.1). Comparative results are presented in Appendix Table A5, showing that the proposed model achieves the highest performance across all baselines on the semi-synthetic data.

### Potential trade-offs among outcomes

In our paper, we consider multiple disease outcomes and identify the optimal drug combination as one that achieves both maximum effectiveness and minimal safety risks. However, there are potential trade-offs among outcomes, particularly in cases where the treatment option with maximum effectiveness may not align with minimal safety risks. To mitigate this, it is essential to incorporate clinicians’ expert knowledge. In real-world clinical settings, clinicians can help determine the relative importance of different outcomes based on their practical experience and model estimated treatment effects. By integrating clinical insights with model outputs, we can carefully assess and prioritize outcomes, ultimately combining them into a single, composite endpoint that balances both effectiveness and safety considerations. This approach allows for the identification of the optimal treatment option based on its estimated effects on the combined outcome, ensuring that the selected treatment best aligns with the patient’s overall needs and preferences.

### Limitations of the Study

We acknowledge that our paper has limitations. First, from a data perspective, the use of observational data presents challenges, primarily due to the absence of ground truths for treatment effects. This lack of true benchmarks makes it difficult to directly evaluate the model’s accuracy in estimating treatment effects. To address this limitation, we proposed the use of a proxy metric, MMIP, to approximate estimation error and supplemented our analysis with experiments on a semi-synthetic dataset that includes simulated ground truths. Second, from methodology perspective, in estimating treatment effects across multiple outcomes and identifying the optimal treatments, potential conflicts among outcomes may arise. We anticipate that, in real clinical settings, incorporating clinicians’ expertise as prior knowledge could help guide the process, minimizing conflicts and enabling convergence toward a reliable recommendation.

## Data Availability

The data we use is from MarketScan Commercial Claims and Encounters (CCAE) and includes approximately 130 million patients from 2012 to 2021. Access to the MarketScan data analyzed in this manuscript is provided by The Ohio State University. The dataset is available at https://www.merative.com/real-world-evidence.

## Acknowledgments

This work was funded in part by the National Institutes of Health (NIH) under award number R01GM141279. The content is solely the responsibility of the authors and does not necessarily represent the official views of the NIH.

## Author contributions

PZ conceived the project. RL and PZ developed the method. RL conducted the experiments. RL, PZ, and LL analyzed the results. RL and PZ wrote the manuscript. All authors read and approved the final manuscript.

## Resource availability

## Lead contact

Further information and requests for resources should be directed to and will be fulfilled by the lead contact, Dr. Ping Zhang.

## Data and code availability

The data we use is from MarketScan Commercial Claims and Encounters (CCAE) and includes approximately 130 million patients from 2012 to 2021. Access to the MarketScan data analyzed in this manuscript is provided by The Ohio State University. The dataset is available at https://www.merative.com/real-world-evidence. The source code for this paper can be downloaded from the GitHub repository at https://github.com/ruoqi-liu/METO.

## Methods

In this section, we introduce the proposed **METO** framework for treatment effect estimation under multiple treatments and multiple outcomes on observational patient data. Fig. 9 shows an illustration of the proposed framework. Specifically, this framework consists of four modules: patient data encoding, multitreatment modeling, multi-outcome prediction, and treatment recommendation.

**Figure 9:**
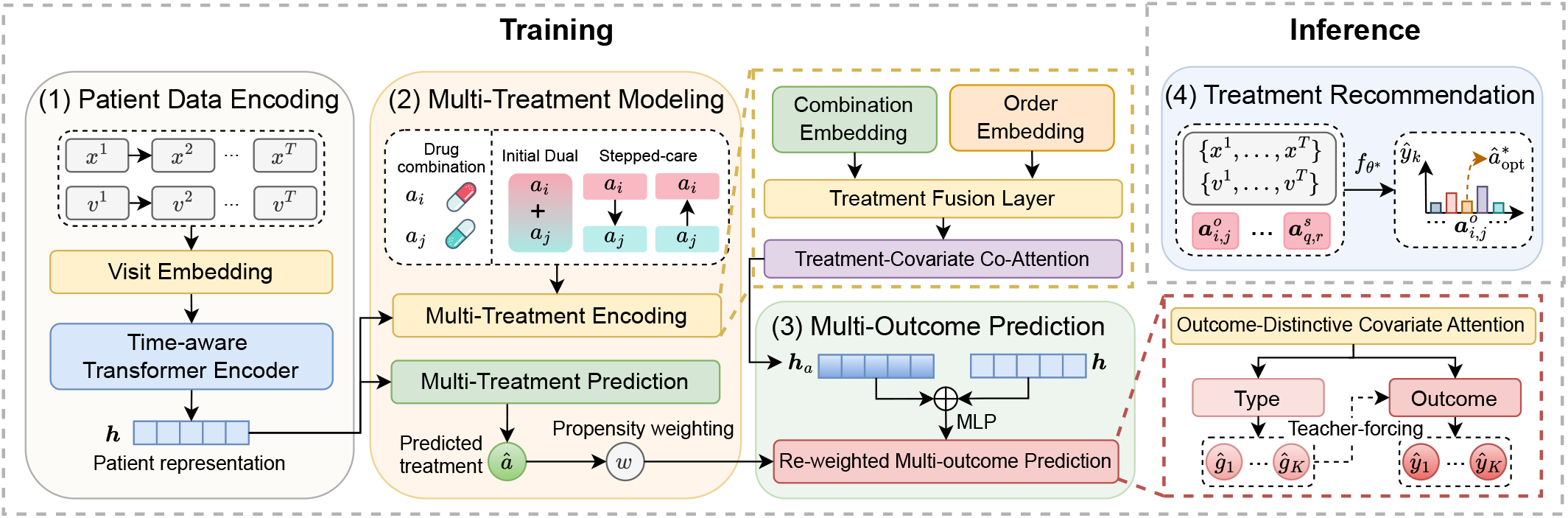
A detailed illustration of **METO**. (1) Patient data encoding: raw patient data is transformed into enriched patient representations. (2) Multi-treatment modeling: drug combinations and administration sequences are encoded, and the probability of receiving each treatment is predicted as balancing scores. (3) Multi-outcome prediction: multiple outcomes are predicted by integrating the patient representations with the treatment representations, re-weighted by the balancing scores. (4) Treatment recommendation: optimal drug combinations are identified based on the estimated treatment effects.

### Preliminaries

### Observational Patient Data

The observational patient data is denoted as 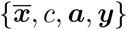. Here, 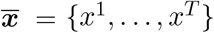 represents the patient’s medical history over T timestamps, capturing the details of each medical visit. The variable c stands for static demographic information, including age (categorical) and gender (binary). Each medical visit, x^*t*^, contains a series of medications and diagnosis codes. Specifically, medications are represented as m_1_,…, m_|ℳ|_ ∈ ℳ, where |ℳ| is the total number of unique medication codes in the dataset. Similarly, diagnosis codes are denoted as d_1_,…, d_|𝒟|_ ∈ 𝒟, with |𝒟| representing the total number of unique diagnosis codes.

### Multiple Treatments

This work studies multiple treatment scenarios for antihypertensive drug combinations. Each drug combination is represented as 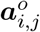, where (a_*i*_, a_*j*_, o) denotes individual drugs a_*i*_, a_*j*_ from the antihypertensive medication set 𝒜, and *o* ∈ 𝒪 of the treatment assignment order. This order includes initial combination therapy (simultaneous administration of a_*i*_ and a_*j*_) and stepped-care protocols (sequential administration of a_*i*_ followed by a_*j*_, or vice versa). Let P denote the total number of unique drug combinations.

### Multiple Outcomes

The investigation extends to multiple outcomes, ***y*** = y_1_,…, y_*K*_, where each outcome y_*k*_, binary in nature, relates to a specific aspect of hypertension disease progression. Here, K denotes the total number of distinct outcomes. Each outcome y_*k*_ is also associated with a type label g_*k*_, categorizing it as either an effectiveness outcome or a safety outcome. This classification provides a comprehensive view of the treatment’s impact, enabling the assessment of treatment effects from different perspectives.

### Treatment Effect Estimation

We extend the Neyman-Rubin potential outcome framework ^31^ to accommodate multiple treatments and multiple outcomes. Given patient pre-treatment covariates ***x̄***,c and distinct treatment combinations 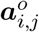 and 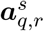 with *i, j, q, r* ∈ 𝒜 and *o, s* ∈ 𝒪, the conditional average treatment effect (CATE) for the k-th outcome is defined as 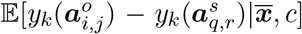. This expression considers 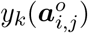 and 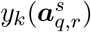 as the potential outcomes under treatment combinations 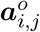 or 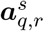, respectively. In observational data, outcomes for the non-received treatments remain unobserved, posing the fundamental causal inference challenge, distinct from traditional supervised learning. To ensure identifiability of treatment effects from observational data, we adhere to three standard assumptions: consistency, positivity, and ignorability ^32^, as elaborated in Appendix A. Under these premises, the treatment effect is estimated as 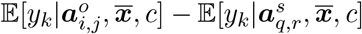.

### Patient Data Encoding

The patient data is originally denoted by high-dimensional medical codes with temporal information. To convert such raw patient data into informative patient representations, we propose patient data encoding, which consists of a visit embedding layer and a time-aware Transformer encoder. Below, we integrate mathematical formulations to elucidate the operations within these components.

### Visit Embedding Layer

The visit embedding layer focuses on the patient’s covariates in medical history, denoted as 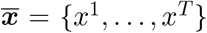. Each element x^*t*^ (where t ∈ 1,…, T) represents the details of a patient’s visit at a specific timestamp. The embedding layer maps these discrete visit records into a continuous vector space, resulting in an embedded representation ***e***(x^*t*^) for each visit. Formally, the embedding of the t-th visit is given by:

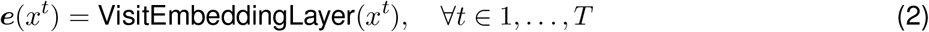

### Time-aware Transformer Encoder

To enhance the patient representation, time information is also encoded and integrated with the visit embeddings. Specifically, the time interval (v_*t*_) between the t-th visit and the initiation of treatment is encoded into a time embedding ***e***(v^*t*^), parallel to the visit embeddings as:

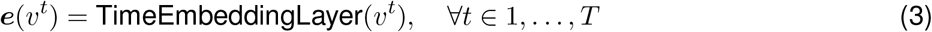

This addition allows the model to account for temporal dynamics explicitly, which are essential in healthcare applications. The combined input to the Transformer encoder ^33^ (see details of Transformer layers in Appendix B.1) consists of both visit and time embeddings. Let ***h***^0^ denote the initial hidden representation as below:

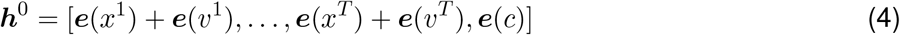

The Transformer processes this enriched input through L layers:

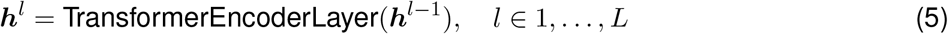

The final output ***h***^*L*^ is a comprehensive representation of the patient, effectively capturing both the temporal aspects of the medical history and static demographic information. This encoded information (denoted as ***h*** for simplicity) is then utilized in subsequent multi-treatment modeling and multi-outcome prediction.

### Multi-Treatment Modeling

This module is designed to address the complexities of multi-treatment modeling, specifically for drug combinations in hypertension management. First, the module encodes multiple drug combinations into latent embeddings. Then, it models the relationships between covariates and treatments to extract treatment-related information. The predicted treatment probability is further utilized as balancing weights to adjust for confounding bias.

### Multi-Treatment Encoding

The initial phase of this module focuses on encoding the drug combinations into an embedding space. Given a drug combination 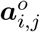, which includes two individual drugs (a_*i*_ and a_*j*_) and their assignment order o, we approach the encoding process in a two-fold manner to capture both the drug-specific information and its sequential information.

Firstly, each drug in the combination, a_*i*_ and a_*j*_, is encoded separately through a drug embedding layer, which transforms the discrete drug identifiers into continuous vector representations. The embeddings for a_*i*_ and a_*j*_ are obtained as:

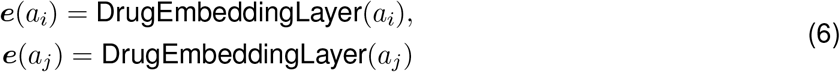

The assignment order o, which indicates whether the drugs are administered simultaneously or sequentially, is also encoded. The order encoding captures the temporal aspect of the treatment administration. This is represented as:

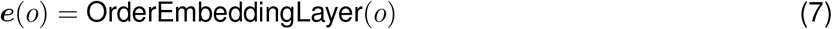

Secondly, to achieve a deep fusion of individual drugs and assignment order for a comprehensive treatment representation, we propose a treatment fusion layer:

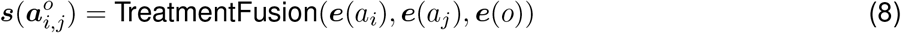

Specifically, this component first concatenates the drug embeddings with the order embedding and then projects this concatenated vector to a latent space through a fully connected layer (FC) for dimensionality alignment and feature extraction:

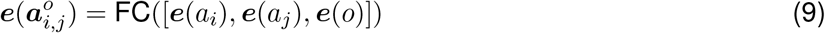

In addition, a self-attention mechanism (multi-head attention mechanism ^33^) is applied to weigh and integrate the information from the drug and order embeddings adaptively:

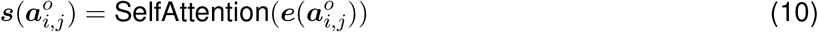

### Treatment-Covariate Co-Attention

This component plays a critical role in modeling the interactions between specific treatments and patient covariates. By considering treatment representation 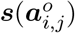 as “queries” and patient hidden representations ***h*** (last hidden state of transformer encoder) as both “keys” and “values”, we employ a co-attention mechanism ^34^ that dynamically adjusts the focus on relevant covariates for each treatment option:

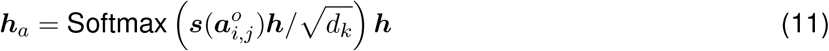

where d_*k*_ is the dimensionality of the key embeddings. This co-attention mechanism allows the model to dynamically focus on different aspects of the patient’s representation in relation to each treatment.

### Treatment Prediction

The pooled patient representation ***h***_*pool*_ (i.e., taking the first `[CLS]` token from ***h***) is processed through a fully-connected layer with a sigmoid activation function *σ* as the last layer to predict the probability of receiving each treatment 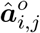 as follows:

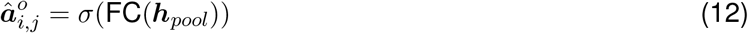

The treatment prediction task is formulated as a multi-class classification problem, where each class corresponds to a specific treatment combination. The treatment prediction loss function is defined as:

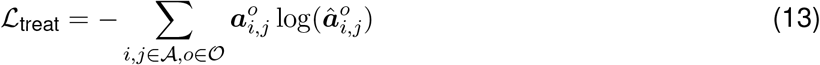

where 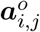 is the ground truth treatment assignment. By minimizing ℒ_treat_, the model is optimized to predict the probability of receiving each treatment combination.

### Propensity Score Weighting

The predicted probability of receiving treatment, also known as the propensity score ^35^, is further leveraged to reduce the bias from confounding factors. Specifically, the balancing weights can be derived from the propensity scores and then used to estimate the potential outcomes. The inverse probability of treatment weighting (IPTW) ^36^ is employed and extended to multiple treatment settings ^37^ as follows,

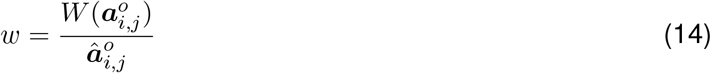

Where 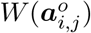 is the marginal probability of treatment 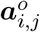 that is included to stabilize the weights ^38^. Then the weight is used in the multi-outcome prediction to re-weight the patient and adjust for confounding bias.

### Multi-Outcome Prediction

This module focuses on predicting multiple disease outcomes in a nuanced and comprehensive manner. This module first encodes the distinctive relationships among different outcomes and patient covariates and then predicts the outcomes, weighted based on the balancing propensity scores.

### Outcome-Distinctive Covariate Attention

This component addresses the distinctive impact that patient covariates have on different disease outcomes. It computes an outcome-distinctive covariate attention score *α* ∈ ℝ^*K×N*^ (where K is the number of outcomes, and N is the length of patient sequence) as follows:

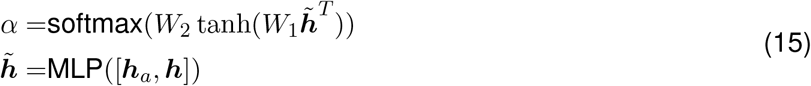

where, 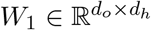 and 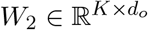 are trainable weight matrices, *α*_*k,n*_ (each element of *α*) indicates the contribution of the n-th covariate to the k-th outcome. Leveraging *α*, the mechanism produces outcome-distinctive patient representations as ***h***_*o*_ = *α****h***, thereby refining the predictions for each distinct outcome.

### Re-weighted Multi-outcome Prediction with Type Information

We recognize the critical role of outcome type (i.e., distinguishing between effectiveness and safety) in accurately predicting patient responses to treatment. These types embody the dual (opposing) aspects of patient outcomes, each critical to understanding the full scope of treatment impacts. To harness this crucial insight, we propose first to predict each outcome type, *ĝ*_*k*_, utilizing this prediction to enhance the subsequent outcome prediction, *ŷ*_*k*_. In a teacher-forcing-like approach, during the training, the model predominantly uses the actual outcome types g_*k*_ as supplementary context for enhancing outcome predictions, while in testing, it relies on its predictive capabilities for *ĝ*_*k*_. Formally, we have

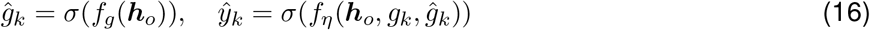

Here, f_*g*_ denotes the function to predict outcome type. f_*η*_ is the function to predict the outcome itself, where η ∈ [0, 1] represents a tunable probability parameter designed to modulate the dependency on true versus predicted outcome types.

To address the potential issue of positive-negative imbalance encountered in outcome prediction (i.e., where the incidence rate of certain disease outcomes may be notably low), we employ the Asymmetric Loss (ASL) ^39^. ASL is adept at handling such imbalances by calculating separate losses for positive and negative samples, denoted as ℒ+ and ℒ*-*, respectively. The formulation is as follows:

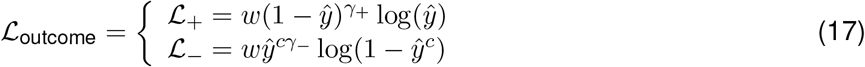

where *ŷ*^*c*^ = max(*ŷ-*c, 0) represents the adjusted outcome probability for negative examples, incorporating a margin c for hard thresholding. The hyperparameters *γ*^−^, *γ*^+^ are set such that *γ*_−_, ≤ *γ*_+_, enabling ASL to appropriately down-weight and apply hard thresholds to easy negative samples, thereby mitigating the imbalances. The balancing weight, w (from Eq. 14), is introduced to adjust the importance of each sample in the loss function based on the treatment assignment probability, which helps to mitigate the bias introduced by the non-random treatment assignment.

### Optimization and Treatment Effect Estimation

The overall loss function integrates the outcome prediction loss −_outcome_ with the treatment prediction loss −_treat_ as:

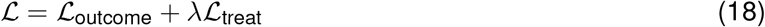

where *λ* is a weighting factor that balances the importance of the outcome prediction loss and the treatment prediction loss. This combined loss function ensures that the model is trained to accurately predict both patient outcomes and treatment assignments, reflecting the complex relations of treatments and their effects.

The estimated treatment effect under a pair of treatments 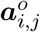 and 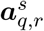 is defined as the differential between the potential outcomes:

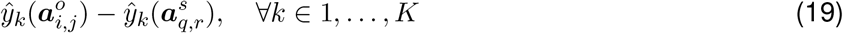

This estimation is essential for elucidating the comparative impact of different treatments, thereby equipping healthcare professionals with the insights needed to make informed treatment selections.

### Treatment Recommendation

This module demonstrates the application of the proposed **METO** in a healthcare problem for antihypertensive drug combination recommendation. Utilizing the estimated treatment effects across diverse drug combinations facilitates the identification of an optimal treatment strategy with maximum therapeutic efficacy and minimum drug safety concerns.

Specifically, the trained model, 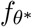, with *θ*^*\**^ representing the optimized parameters, is employed to simultaneously evaluate effectiveness and safety outcomes for each potential treatment regimen concerning a new patient as:

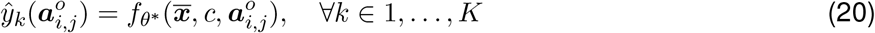

The model evaluates each antihypertensive drug combination 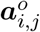 by predicting effectiveness outcomes, 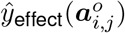, and safety outcomes, 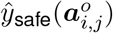. The therapeutic effectiveness improvement over a baseline, denoted as 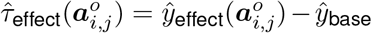, guides the ranking of treatments. The optimal treatment, 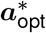, is then identified as the one offering maximal effectiveness improvement while ensuring minimal safety risk:

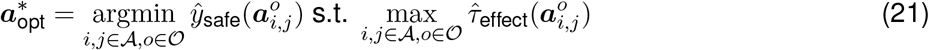

This streamlined approach empowers healthcare professionals to tailor treatment plans precisely, balancing efficacy with safety, to meet individual patient needs effectively.

## Footnotes

1 https://www.merative.com/real-world-evidence

2 https://hcup-us.ahrq.gov/toolssoftware/ccs/ccs.jsp

