## Supplementary for "Estimating the Treatment Effects of Multiple Drug Combinations on Multiple Outcomes in Hypertension"

### Appendix

#### A Causal Assumption

Our study is based on three standard assumptions in causal inference. We elaborate each of them as below:

**Assumption 1 (Consistency)** *The potential outcome under the treatment $A$ equals the observed outcome if the actual treatment is $A$.*

**Assumption 2 (Positivity)** *Given the observational patient covariates $X$, the probability of receiving treatment $A$ is positive, i.e., $0 < P(A|X) < 1, \forall A$ and $X$.*

**Assumption 3 (Strong Ignorability)** *Given the observational patient covariates $X$, the treatment assignment is independent of the $K$ potential outcomes, i.e., $Y_1(A), \ldots, Y_K(A) \perp\!\!\!\perp A|X$.*

Assumption 1 is fundamental to the potential outcome framework[31], which defines factual outcomes, counterfactual outcomes, and treatment effects. The assumption requires that the treatment specified in the study is precise enough that any variation within the treatment specification will not lead to a different outcome. Assumption 2 suggests that all patients, regardless of their covariates, have the potential to receive the treatment. Without this assumption, it would be impossible to derive counterfactuals for patients who do not have any chance of being in other treatment groups. Assumption 3 states that potential outcomes are independent of the treatment assignment, given the set of observed covariates.

#### B Methodology

##### B.1 Transformer Architecture

The Transformer architecture plays a crucial role in our approach due to its ability to handle sequence data effectively. This is particularly relevant in analyzing patient data where temporal patterns can provide significant insights. Each single Transformer encoder block consists of a multi-head self-attention layer followed by a fully-connected feed-forward layer[33]. The multi-head attention is the most crucial part which can be calculated as:

$$\begin{aligned} \text{MultiHead}(\boldsymbol{h}) &= \text{Concat}(\text{head}_1, \ldots, \text{head}_h)W^O, \\ \text{head}_i &= \text{Attention}(\boldsymbol{h}W_i^Q, \boldsymbol{h}W_i^K, \boldsymbol{h}W_i^V), \\ \text{Attention}(Q, K, V) &= \text{Softmax}(\frac{QK^T}{\sqrt{d}})V, \end{aligned} \tag{1}$$

where $\boldsymbol{h} \in \mathbb{R}^{d \times d_{\text{model}}}$ denotes the hidden representations and $d$ is the input sequence length. $W_i^Q \in \mathbb{R}^{d_{\text{model}} \times d}$, $W_i^K \in \mathbb{R}^{d_{\text{model}} \times d}$, $W_i^V \in \mathbb{R}^{d_{\text{model}} \times d}$, $W^O \in \mathbb{R}^{nd \times d_{\text{intermediate}}}$ are learnable parameter matrices. $d = d_{\text{model}}/n$ and $n$ is the number of attention heads.

##### B.2 Patient Embedding

We encode the patient history (medications and diagnosis as medical codes, and type information) into embeddings through linear embedding layers. Mathematically, we have

$$e(x^t) = W_{\text{emb}}^{\text{code}} m_{\text{code}} + W_{\text{emb}}^{\text{type}} m_{\text{type}}, \tag{2}$$

where $W_{\text{emb}}^{\text{code}} \in \mathbb{R}^{d_{\text{emb}} \times d_{\text{code}}}$, $W_{\text{emb}}^{\text{type}} \in \mathbb{R}^{d_{\text{emb}} \times d_{\text{type}}}$, $d_{\text{emb}}$ is the embedding dimension, $d_{\text{code}}$ is the total number of medications and diagnosis, $m_{\text{code}} \in \mathbb{R}^{d_{\text{code}}}$ is the collection of medications and diagnosis, and $m_{\text{type}} \in \mathbb{R}^{d_{\text{type}}}$ is the covariate type to distinguish among medication, diagnosis, and demographics. We encode the time interval $v^t$ between the $t$-th visit and the initiation of treatment into a time embedding. Mathematically, we have

$$e(v^t) = W_{\text{emb}}^{\text{time}} m_{\text{time}}, \tag{3}$$

where $W_{\text{emb}}^{\text{time}} \in \mathbb{R}^{d_{\text{emb}} \times d_{\text{time}}}$, $d_{\text{time}}$ is the maximal length of visit time, which is defined as $T/\delta$, and $\delta$ is a fixed window length used to discretize the continuous time interval. $m_{\text{time}} \in \mathbb{R}^{d_{\text{time}}}$ denotes the time interval between the visit and treatment index date divided by the time window $\delta$. We encode demographics $c$ as

$$e(c) = W_{\text{emb}}^{\text{demo}} m_{\text{demo}}, \tag{4}$$

where $d_{\text{demo}}$ is the feature space for demographics (all age groups and genders), and $m_{\text{demo}}$ is the current age group

### C Additional Experimental Setup

#### C.1 Dataset

The definition of hypertension using ICD codes is provided in Table A1. The studied first-line antihypertensive drug ingredients are detailed in Table A2. The definition of all six outcomes using ICD codes are provided in Table A3.

Table A1: The definition of hypertension in observational health data.

| | |
|---|---|
| Diagnosis | ICD-9 codes:<br>401-405 Hypertensive disease,<br>995.1 Angioneurotic edema not elsewhere classified<br>ICD 10 codes:<br>I10-I1A Hypertensive diseases |

Table A2: First-line antihypertensive drug ingredients in four major drug classes.

| TZDs | | ACEIs | | ARBs | | CCBs | |
|---|---|---|---|---|---|---|---|
| Drug | RxNorm | Drug | RxNorm | Drug | RxNorm | Drug | RxNorm |
| Chlorthalidone | 2409 | Benazepril | 18867 | Azilsartan | 1091643 | Amlodipine | 17767 |
| Hydrochlorothiazide | 5487 | Celiprolol | 20498 | Candesartan | 214354 | Felodipine | 4316 |
| Indapamide | 5764 | Enalapril | 3827 | Eprosartan | 83515 | Isradipine | 33910 |
| Metolazone | 6916 | Fosinopril | 50166 | Irbesartan | 83818 | Nicardipine | 7396 |
| | | Lisinopril | 29046 | Losartan | 52175 | Nifedipine | 7417 |
| | | Moexipril | 30131 | Olmesartan | 321064 | Nisoldipine | 7435 |
| | | Perindopril | 54552 | Telmisartan | 73494 | Diltiazem | 3443 |
| | | Quinapril | 35208 | Valsartan | 69749 | Verapamil | 11170 |
| | | Ramipril | 35296 | | | | |
| | | Trandolapril | 38454 | | | | |

#### C.2 Evaluation Metrics

Our evaluation metric for TEE is based on a recent proxy metric, called influence function-based precision of estimating heterogeneous effects (IF-PEHE)[24]. We elaborate the computation procedure of IF-PEHE as follows:

- Step 1: Train two XGBoost[40] classifiers for potential outcome prediction denoted by $\mu_0$ and $\mu_1$, where $\mu_a = P(y(a) = 1|X = x)$ using the training set $\mathcal{Z}_{\text{train}}$. Then calculate the plug-in estimation $\widetilde{T} = \mu_1 - \mu_0$ Train an XGBoost[40] classifier propensity score function (i.e., the probability of receiving treatment) $\widetilde{\pi} = P(a = 1|X = x)$.

Table A3: The definition of six outcomes in observational health data.

| | |
|---|---|
| Stroke | ICD-9 codes:<br>430 Subarachnoid hemorrhage,<br>431 Intracerebral hemorrhage,<br>432 Other and unspecified intracranial hemorrhage,<br>433.11 Occlusion and stenosis of carotid artery with cerebral infarction,<br>433.21 Occlusion and stenosis of vertebral artery with cerebral infarction,<br>433.31 Occlusion and stenosis of multiple and bilateral precerebral arteries with cerebral infarction,<br>433.81 Occlusion and stenosis of other specified precerebral artery with cerebral infarction,<br>433.91 Occlusion and stenosis of unspecified precerebral artery with cerebral infarction,<br>434.01 Cerebral thrombosis with cerebral infarction,<br>434.11 Cerebral embolism with cerebral infarction,<br>434.91 Cerebral artery occlusion, unspecified with cerebral infarction,<br>436 Acute, but ill-defined, cerebrovascular disease<br>ICD 10 codes:<br>I60 subarachnoid hemorrhage,<br>I61 intracerebral hemorrhage,<br>I63 cerebral infarction |
| Myocardial infarction | ICD-9 codes:<br>410 Acute myocardial infarction<br>ICD 10 codes:<br>I21 Acute myocardial infarction,<br>I22 Subsequent ST elevation (STEMI) and non-ST elevation (NSTEMI) myocardial infarction |
| Heart failure | ICD-9 codes:<br>428 Heart failure,<br>425 Cardiomyopathy<br>ICD 10 codes:<br>I50 Heart failure,<br>I42 Cardiomyopathy |
| Acute kidney failure | ICD-9 codes:<br>584 Acute kidney failure<br>ICD 10 codes:<br>N17 Acute kidney failure |
| Gout | ICD-9 codes:<br>274 Gout<br>ICD 10 codes:<br>M10 Gout,<br>M1A Chronic gout |
| Venous thromboembolism | ICD-9 codes:<br>415.1 Pulmonary embolism and infarction,<br>451 Phlebitis and thrombophlebitis,<br>453 Other venous embolism and thrombosis<br>ICD 10 codes:<br>I80 Phlebitis and thrombophlebitis,<br>O87.0 Superficial thrombophlebitis in the puerperium,<br>O87.1 Deep phlebothrombosis in the puerperium,<br>I26 Pulmonary embolism |

- Step 2: Given the estimated treatment effect $\hat{T}(x_i)$ on the test set $\mathcal{Z}_{\text{test}}$, calculate the IF-PEHE with the influence function $\hat{l}$ as:

$$\text{IF-PEHE} = \sum_{x_i \in \mathcal{Z}_{\text{test}}} [(\hat{T}(x_i) - \widetilde{T}(x_i))^2 + \hat{l}(x_i)],$$

$$\hat{l}(x) = (1-B)\widetilde{T}^2(x) + (\widetilde{T}(x) - \hat{T}(x)) - W(\widetilde{T}(x) - \hat{T}(x))^2 + \hat{T}^2(x),$$

where $W = (a - \widetilde{\pi}(x))$, $B = 2a(a - \widetilde{\pi}(x))C^{-1}$, $C = \widetilde{\pi}(x)(1 - \widetilde{\pi}(x))$.

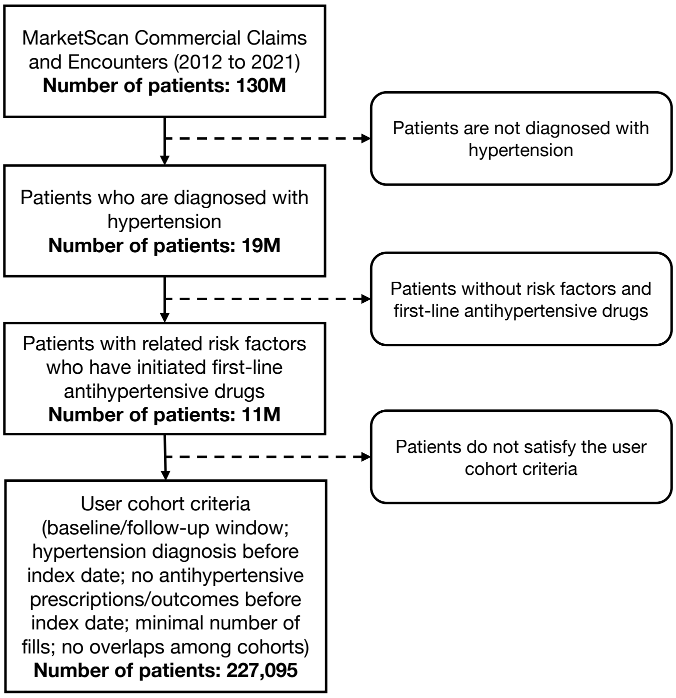


Figure A1: The flowchart of the user cohort selection. We start from a total of 130M patients in the MarketScan Commercial Claims and Encounters database and filter out the studied user cohort of 227K patients, including all patients with hypertension who initiated first-line antihypertensive drug combinations and satisfied the user cohort selection criteria.

##### C.3 Implementation Details

The training process is conducted on 4 NVIDIA GeForce RTX 2080 Ti 16GB GPUs. We use the Adam optimizer given its ability to adapt the learning rate for each parameter, which can lead to more efficient and robust model training. We tune the model using a range of parameter configurations for the Transformer and find that a 12-layer Transformer with 768 hidden units, 12 attention heads and 3072 intermediate size yields the best performance. Detailed hyperparameter tuning is shown in Table A4.

Table A4: The search space of hyperparameters and the optimal parameters utilized during the model training.

| Parameters | Search Space | Optimal Value |
|---|---|---|
| Maximum Epochs | {1, 5,10,15,20} | 10 |
| Initial Learning Rate | {1e-5, 3e-5, 5e-5} | 5e-5 |
| Batch Size | {8, 16, 32, 64} | 32 |
| Sequence Length | 256 | 256 |
| Fixed Window Length | 30 | 30 |
| Baseline Window | {90, 180, 360, 720} | 360 |
| Dropout | 0.1 | 0.1 |
| $\beta$ | {0, 0.2, 0.4, 0.6, 0.8, 1} | 0.6 |
| $\lambda$ | {0, 0.2, 0.4, 0.6, 0.8, 1} | 1 |

### D Additional Results

#### D.1 Results on semi-synthetic dataset

We create a semi-synthetic dataset based on real patient data obtained from the MarketScan data. Specifically, we simulate treatment assignments $\boldsymbol{a}_{i,j}^{o}$ and potential outcomes $\boldsymbol{y} = \{y_1, \dots, y_K\}$ using pre-treatment covariates $\boldsymbol{x}$ (i.e., historical co-medication, co-morbidities, and demographics). The treatment assignment is simulated by $\boldsymbol{a}_{i,j}^{o}|\overline{x} \sim$ Categorical$(\pi(\overline{x}))$, where $\pi(\overline{x}) =$ Softmax$(S^T\overline{x} + m)$, $S \sim \mathcal{N}(0^{|\mathcal{V}|\times|\mathcal{T}|}, 0.1 \cdot I)$, $|\mathcal{V}|$ is the cardinality of the medical feature vocabulary, $|\mathcal{T}|$ is the number of possible treatment combinations, $m \sim \mathcal{N}(0^{|\mathcal{T}|}, 0.1 \cdot I)$, and $\overline{x}$ denotes the aggregation of all historical covariates. The outcomes are simulated by $y_k|\overline{x}, \boldsymbol{a}_{i,j}^{o} \sim$ Bernoulli(Sigmoid$(w_k^T\overline{x} + \beta_k^T\boldsymbol{a}_{i,j}^{o} + n_k)$), where $w_k \sim \mathcal{N}(0^{|\mathcal{V}|}, 0.1 \cdot I)$, $\beta_k \sim \mathcal{N}(0^{|\mathcal{T}|}, 1 \cdot I)$ is the treatment effect parameter vector for outcome $y_k$, $\boldsymbol{a}_{i,j}^{o}$ is represented as a one-hot encoded vector indicating the treatment combination and order, $n_k \sim \mathcal{N}(0, 0.1)$, and $k = 1, \dots, K$.

Table A5 shows the comparison results on the semi-synthetic dataset. We observe that the proposed model yields the best performance among all the baselines.

Table A5: Comparison with state-of-the-art methods on semi-synthetic datasets. The precision of Estimating Heterogeneous Effects (PEHE) is reported. The results are the average and standard deviation over 20 runs.

| | Stroke | MI | HF | AFK | Gout | VTE |
|---|---|---|---|---|---|---|
| S-learner | 0.578 ± 0.027 | 0.553 ± 0.028 | 0.554 ± 0.026 | 0.568 ± 0.027 | 0.550 ± 0.011 | 0.567 ± 0.015 |
| T-learner | 0.551 ± 0.015 | 0.558 ± 0.018 | 0.559 ± 0.020 | 0.552 ± 0.016 | 0.561 ± 0.019 | 0.563 ± 0.021 |
| TARNet | 0.462 ± 0.018 | 0.498 ± 0.022 | 0.487 ± 0.018 | 0.472 ± 0.014 | 0.475 ± 0.017 | 0.461 ± 0.024 |
| DragonNet | 0.486 ± 0.025 | 0.492 ± 0.024 | 0.452 ± 0.012 | 0.481 ± 0.014 | 0.495 ± 0.020 | 0.459 ± 0.027 |
| PerfectMatch | 0.385 ± 0.011 | 0.442 ± 0.014 | 0.417 ± 0.027 | 0.423 ± 0.025 | 0.428 ± 0.021 | 0.431 ± 0.028 |
| MEMENTO | 0.424 ± 0.019 | 0.388 ± 0.020 | 0.432 ± 0.020 | 0.381 ± 0.013 | 0.38 ± 0.021 | 0.446 ± 0.021 |
| TECE-VAE | 0.420 ± 0.028 | 0.437 ± 0.012 | 0.386 ± 0.011 | 0.408 ± 0.017 | 0.420 ± 0.023 | 0.448 ± 0.017 |
| LR-learner | 0.407 ± 0.017 | 0.423 ± 0.016 | 0.386 ± 0.013 | 0.429 ± 0.017 | 0.431 ± 0.021 | 0.440 ± 0.024 |
| TransTEE | 0.376 ± 0.026 | 0.381 ± 0.017 | 0.366 ± 0.025 | 0.370 ± 0.017 | 0.353 ± 0.015 | 0.399 ± 0.023 |
| **METO** | **0.320 ± 0.018** | **0.332 ± 0.012** | **0.308 ± 0.022** | **0.323 ± 0.016** | **0.311 ± 0.014** | **0.344 ± 0.014** |

#### D.2 Results on real-world dataset

We provide the full results of performance comparison in Table A6.

#### D.3 Ablation study on propensity score weighting

We conducted an ablation study on propensity score weighting by excluding the weighting module from the treatment effect estimation. Table A7 shows that our model (**METO**) with weighting outperforms the variant without weighting (w/o PW).

#### D.4 Covariate balancing

As shown in Fig. A2, the weighted ASMD for all covariates falls below the threshold and is smaller than the unweighted ASMD, demonstrating effective covariate balance.

Table A6: Performance comparison for factual outcome prediction (AUPR) and treatment effect estimation (MM-IP: IF-PEHE for multiple treatments and multiple outcomes). The results are averaged over 20 random runs.

| Method | Stroke | | MI | | HF | |
|---|---|---|---|---|---|---|
| | AUPR ↑ | MM-IP ↓ | AUPR ↑ | MM-IP ↓ | AUPR ↑ | MM-IP ↓ |
| S-learner | 0.551 ± 0.010 | 0.284 ± 0.005 | 0.188 ± 0.008 | 0.355 ± 0.006 | 0.329 ± 0.012 | 0.378 ± 0.009 |
| T-learner | 0.544 ± 0.007 | 0.297 ± 0.009 | 0.181 ± 0.009 | 0.360 ± 0.005 | 0.325 ± 0.011 | 0.386 ± 0.006 |
| TARNet | 0.569 ± 0.009 | 0.271 ± 0.008 | 0.196 ± 0.010 | 0.351 ± 0.005 | 0.342 ± 0.010 | 0.357 ± 0.008 |
| DragonNet | 0.574 ± 0.004 | 0.262 ± 0.012 | 0.198 ± 0.009 | 0.344 ± 0.004 | 0.351 ± 0.011 | 0.338 ± 0.012 |
| PerfectMatch | 0.591 ± 0.008 | 0.239 ± 0.007 | 0.202 ± 0.009 | 0.361 ± 0.005 | 0.388 ± 0.012 | 0.312 ± 0.004 |
| MEMENTO | 0.584 ± 0.011 | 0.244 ± 0.008 | 0.208 ± 0.009 | 0.354 ± 0.005 | 0.382 ± 0.012 | 0.317 ± 0.009 |
| TECE-VAE | 0.605 ± 0.007 | 0.230 ± 0.003 | 0.211 ± 0.008 | 0.355 ± 0.005 | 0.397 ± 0.013 | 0.302 ± 0.007 |
| LR-learner | 0.609 ± 0.006 | 0.225 ± 0.010 | 0.206 ± 0.010 | 0.364 ± 0.006 | 0.392 ± 0.011 | 0.308 ± 0.004 |
| TransTEE | 0.617 ± 0.009 | 0.221 ± 0.005 | 0.235 ± 0.007 | 0.321 ± 0.005 | 0.409 ± 0.013 | 0.284 ± 0.008 |
| NCoRE | 0.577 ± 0.008 | 0.249 ± 0.004 | 0.207 ± 0.010 | 0.361 ± 0.006 | 0.388 ± 0.012 | 0.313 ± 0.013 |
| **METO** | **0.656 ± 0.011** | **0.185 ± 0.003** | **0.329 ± 0.009** | **0.225 ± 0.005** | **0.442 ± 0.013** | **0.236 ± 0.004** |

| Method | AKF | | Gout | | VTE | |
|---|---|---|---|---|---|---|
| | AUPR ↑ | MM-IP ↓ | AUPR ↑ | MM-IP ↓ | AUPR ↑ | MM-IP ↓ |
| S-learner | 0.218 ± 0.013 | 0.370 ± 0.007 | 0.339 ± 0.008 | 0.325 ± 0.008 | 0.274 ± 0.007 | 0.356 ± 0.008 |
| T-learner | 0.212 ± 0.009 | 0.377 ± 0.007 | 0.331 ± 0.009 | 0.332 ± 0.005 | 0.267 ± 0.008 | 0.363 ± 0.004 |
| TARNet | 0.233 ± 0.011 | 0.359 ± 0.006 | 0.349 ± 0.007 | 0.318 ± 0.011 | 0.288 ± 0.009 | 0.355 ± 0.007 |
| DragonNet | 0.251 ± 0.010 | 0.342 ± 0.006 | 0.356 ± 0.008 | 0.303 ± 0.009 | 0.294 ± 0.008 | 0.332 ± 0.006 |
| PerfectMatch | 0.274 ± 0.008 | 0.331 ± 0.006 | 0.378 ± 0.007 | 0.289 ± 0.005 | 0.312 ± 0.008 | 0.317 ± 0.004 |
| MEMENTO | 0.270 ± 0.007 | 0.338 ± 0.006 | 0.372 ± 0.008 | 0.294 ± 0.007 | 0.310 ± 0.008 | 0.322 ± 0.009 |
| TECE-VAE | 0.290 ± 0.009 | 0.325 ± 0.007 | 0.381 ± 0.007 | 0.283 ± 0.005 | 0.319 ± 0.008 | 0.309 ± 0.007 |
| LR-learner | 0.296 ± 0.006 | 0.321 ± 0.007 | 0.385 ± 0.007 | 0.280 ± 0.012 | 0.326 ± 0.007 | 0.315 ± 0.004 |
| TransTEE | 0.303 ± 0.012 | 0.311 ± 0.006 | 0.403 ± 0.006 | 0.261 ± 0.007 | 0.338 ± 0.007 | 0.276 ± 0.011 |
| NCoRE | 0.275 ± 0.008 | 0.334 ± 0.007 | 0.370 ± 0.008 | 0.297 ± 0.009 | 0.311 ± 0.007 | 0.326 ± 0.006 |
| **METO** | **0.364 ± 0.014** | **0.238 ± 0.007** | **0.453 ± 0.007** | **0.204 ± 0.006** | **0.363 ± 0.008** | **0.201 ± 0.003** |

Table A7: Ablation study on propensity score weighting

| Method | Stroke | | MI | | HF | | AKF | | Gout | | VTE | |
|---|---|---|---|---|---|---|---|---|---|---|---|---|
| | AUPR ↑ | MM-IP ↓ | AUPR ↑ | MM-IP ↓ | AUPR ↑ | MM-IP ↓ | AUPR ↑ | MM-IP ↓ | AUPR ↑ | MM-IP ↓ | AUPR ↑ | MM-IP ↓ |
| w/o PW | 0.622 | 0.216 | 0.272 | 0.288 | 0.413 | 0.265 | 0.327 | 0.277 | 0.426 | 0.233 | 0.341 | 0.249 |
| **METO** | **0.656** | **0.185** | **0.329** | **0.225** | **0.442** | **0.236** | **0.364** | **0.238** | **0.453** | **0.204** | **0.363** | **0.201** |

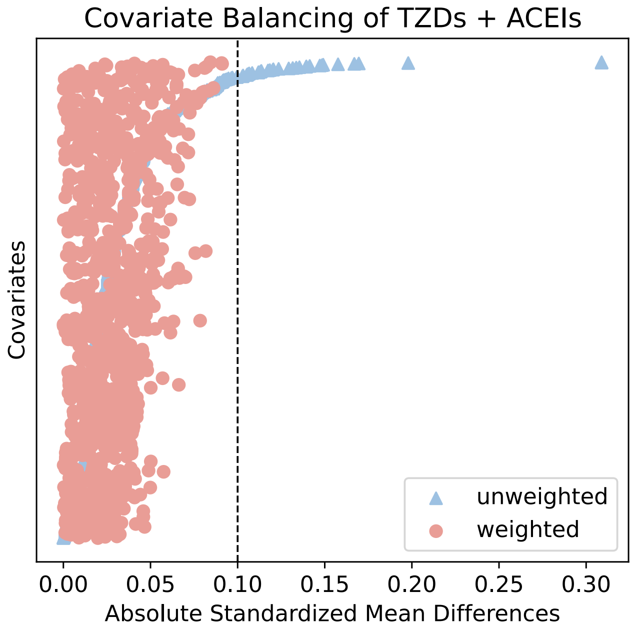


Figure A2: Performance of covariate balancing of TZDs and ACEIs. The absolute standard mean difference (ASMD) values of all covariates before and after weighting are presented. The covariates are sorted by their ASMD in the original unweighted data. The vertical dotted red line at 0.10 represents the threshold for balance, with points closer to zero indicating better balance.
